# Development and retrospective validation of SCOUT: scalable clinical oversight of large language models via uncertainty triangulation

**DOI:** 10.64898/2026.02.08.26345860

**Authors:** Zhengqing Ba, Ming He, Haorui He, Qingan Fu, Jingrong Lai, Ruiqi Zhang, Xiaolin Diao, Mengyuan Liu, Zuoxiang Wang, Ximei Wang, Sheng Zhao, Yalin Zhu, Henghongyu Chen, Yuanwang Qiu, Qilin Su, Jinghang Xu, Fenghuan Hu, Xinlin Luo, Hongwu Chen, Mingqi Zheng, Bing Xu, Jiabao Liu, Ning Guo, Xiaojin Gao, Guiqiang Wang, Yongjian Wu

## Abstract

Large language models (LLMs) are increasingly used in clinical workflows, yet requiring clinician review of every AI output negates the efficiency gains that motivate their adoption. We present SCOUT (Scalable Clinical Oversight via Uncertainty Triangulation), a model-agnostic meta-verification framework that selectively defers unreliable LLM predictions to clinicians by triangulating three orthogonal signals: model heterogeneity, stochastic inconsistency, and reasoning critique. In this retrospective development and validation study, we derived the framework on a discovery cohort (n = 405) and validated it across three clinically distinct tasks using 4 independent retrospective cohorts: coronary heart disease subtyping (n = 2,271), liver cancer screening from radiology reports (n = 3,373), and diseased coronary vessel counting (n = 286). SCOUT reduced the volume of cases requiring human review by 45% to 83%, with projected final accuracy of 99.1% to 100.0% assuming expert correction of all flagged cases. SCOUT provides a scalable, retrospectively validated approach for deploying generative AI in clinical medicine without compromising patient safety. Prospective randomized validation is underway to confirm real-world clinical utility.

## Introduction

Large language models (LLMs) are rapidly moving from experimental prototypes to operational tools in clinical care. In a randomized trial of 238 outpatient physicians, LLM-powered documentation reduced physician time-in-note by up to 9.5% and improved burnout metrics across 14 specialties^1^. A hybrid deep-learning–LLM system improved diabetes control and medication adherence^2^. High-performance open-weight models have further accelerated this trend: within months of its public release, DeepSeek was integrated into over 300 hospitals in China for clinical decision support and patient communication^3^, with diagnostic accuracy approaching that of leading proprietary models^4^. However, adoption is outpacing governance. Many deployments remain weakly supervised^5^, and LLMs can produce fluent but factually incorrect outputs, often driven by reasoning breakdowns rather than knowledge gaps, a failure mode that can directly lead to incorrect diagnosis and treatment^6^.

The instinctive response is to require clinician review of every AI output^7,8^, but if every output must be re-read, the productivity gains that motivate adoption evaporate, creating a fundamental efficiency– safety paradox. Compounding this problem, perpetual vigilance is cognitively unsustainable. As systems appear reliable in the majority of cases, clinicians drift toward automation bias, turning what was meant as a safety check into a rubber stamp^9,10^. A more scalable alternative is selective deferral, routing only the cases most likely to be wrong to clinician review^11,12^ while allowing low-risk outputs to pass with minimal friction^13,14^.

Selective deferral is well established for discriminative imaging models. The CoDoC framework, for instance, reduced false positives by 25% in mammography screening while cutting reader workload by 66% through complementarity-driven deferral^15^. However, these approaches require the model to output numerical confidence scores and labeled task-specific data to train a deferral rule^16^, requirements that are difficult to meet in the generative setting. Generative LLMs compound the challenge on three fronts. First, LLMs’ self-reported confidence is unreliable. They can express certainty even when their reasoning is flawed^17-19^. Second, naive threshold-based routing ignores clinician cognitive state and risks alert fatigue^20,21^. Third, training dedicated supervisor models demands substantial annotation that rarely transfers across tasks or proprietary deployments^13,16,22^. Together, these constraints point to a practical need for a model-agnostic audit layer capable of flagging risky outputs without access to the model’s internal confidence or task-specific training data.

Here we present SCOUT (Scalable Clinical Oversight via Uncertainty Triangulation), an algorithmic meta-verification framework designed to resolve this paradox (Fig. 1). Rather than relying on internal model logits, SCOUT implements selective deferral by triangulating three orthogonal external signals: whether a different LLM agrees (model heterogeneity), whether the same model answers consistently on repeated runs (stochastic inconsistency), and whether the reasoning contains logical flaws (reasoning critiques). In this study, we describe the development and retrospective validation of SCOUT across three distinct clinical tasks involving 5,930 cases from 4 independent retrospective data sources, spanning diagnostic reasoning, imaging-based screening, and structured information extraction.

**Fig. 1.**
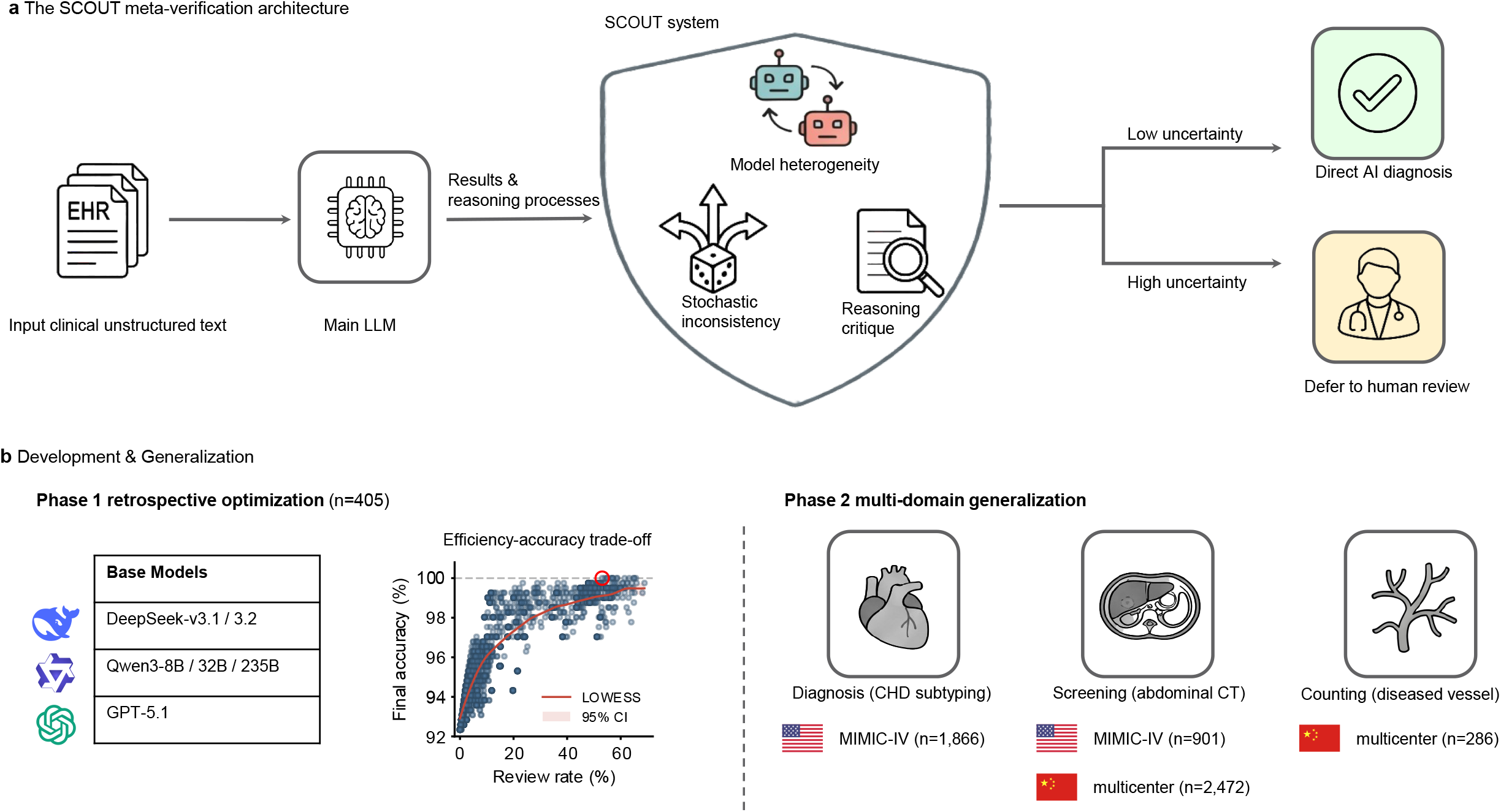
The SCOUT meta-verification framework and study design. **a**, Schematic of the SCOUT architecture. Unstructured clinical text is processed by a main large language model (*M*main) to generate a diagnostic prediction with chain-of-thought reasoning. The prediction is then evaluated by three orthogonal verification strategies—model heterogeneity (S1), stochastic inconsistency (S2) and reasoning critique (S3)—whose outputs are combined into a binary deferral function *D*(*x*). Cases classified as high uncertainty (*D*(*x*) = 1) are routed to human review; low-uncertainty cases (*D*(*x*) = 0) are accepted as direct AI diagnoses. **b**, Development and validation pipeline. Phase 1 (left): retrospective Pareto optimization on a discovery cohort identified the configuration that maximized final accuracy while minimizing the physician review rate, specifically for the DeepSeek-V3.1 chain-of-thought model. Phase 2 (right): the locked configuration was evaluated across three clinically distinct tasks—diagnostic reasoning (coronary heart disease subtyping), imaging screening (liver cancer risk detection) and information extraction (vessel counting)—using Chinese multicentre and US MIMIC-IV datasets.

## Results

### Efficacy of individual and combined verification strategies

We evaluated three orthogonal uncertainty triangulation strategies — model heterogeneity (S_1_), stochastic inconsistency (S_2_), and reasoning critique (S_3_) — across five large language models (LLMs) with standalone accuracies of 90% or above (Extended Data Table 1). Using linear mixed-effects models to account for model architecture as a random effect, all three strategies independently concentrated errors in the deferred subset at rates significantly exceeding the random-deferral baseline (efficiency ratio > 1.0; all P < 0.001; Fig. 2).

**Fig. 2.**
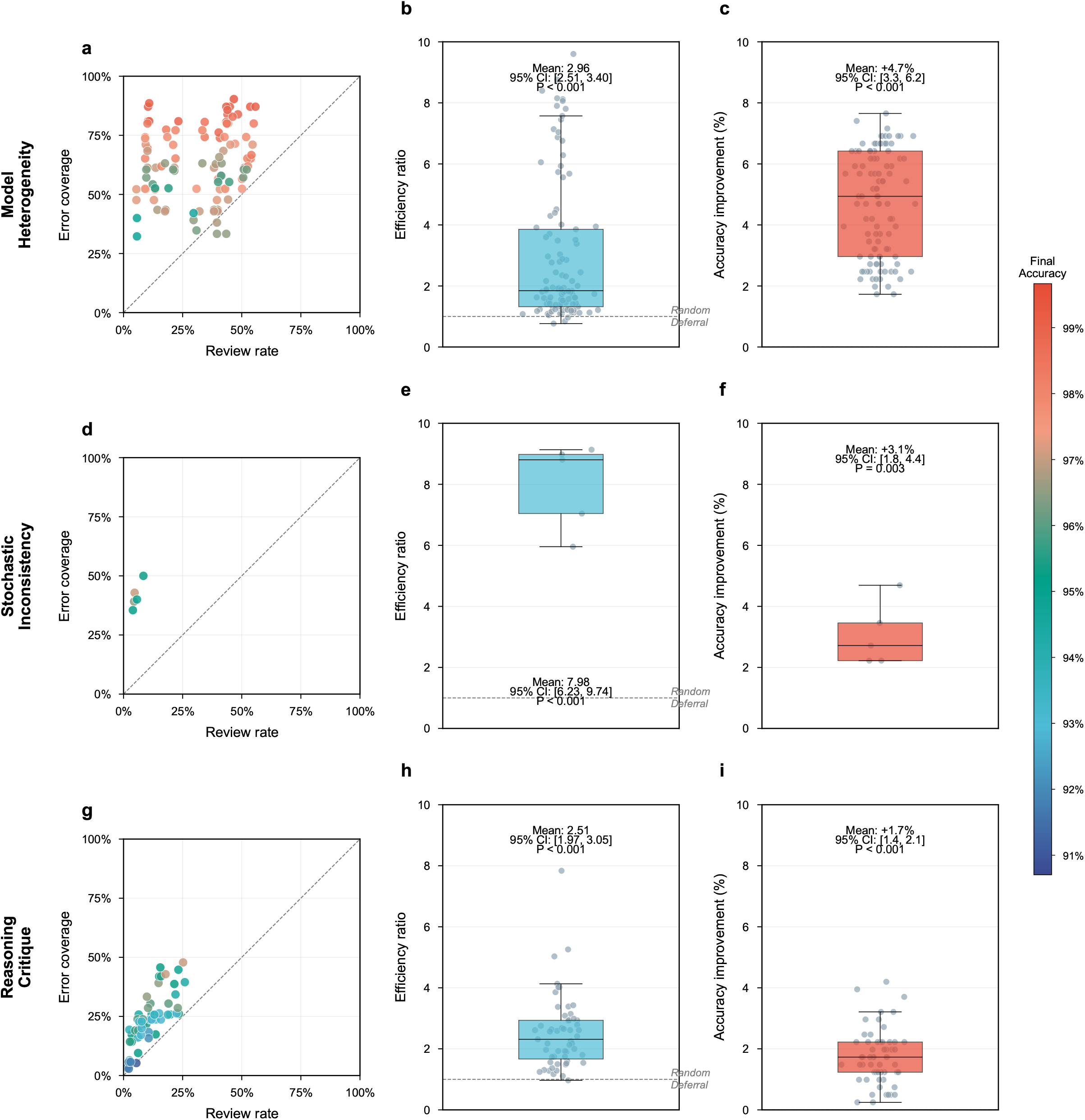
Efficacy of individual uncertainty triangulation strategies in the discovery cohort. Performance of the three verification strategies across 22 model configurations (*n* = 405 cases). **a**,**d**,**g**, Cost– benefit landscapes. The *y* axis represents error coverage and the *x* axis represents the review rate. The diagonal dashed line indicates the random-deferral baseline (efficiency ratio = 1.0); point colours denote final system accuracy. **b**,**e**,**h**, Efficiency ratio (ER = error coverage / review rate) distributions. The dashed line at *y* = 1 marks the random-selection threshold. **c**,**f**,**i**, Absolute accuracy improvement over the AI-only baseline (*y* = 0). Box plots show medians (centre lines) and interquartile ranges (box limits); whiskers extend to 1.5× IQR. Individual data points are overlaid. Means, 95% CIs and *P* values were estimated using linear mixed-effects models with model architecture as a random effect for hierarchical strategies (S1 and S3) and one-sample *t*-tests for stochastic sampling (S2). All tests were two-sided.

The three strategies captured complementary failure modes. S_1_ yielded the largest absolute gain in final accuracy (mean +4.7 percentage points, 95% CI 3.3–6.2, P < 0.001; Fig. 2a–c), reflecting its capacity to break model-specific reasoning blind spots through architectural diversity. S_2_ was the most efficient signal, capturing errors at nearly eight times the rate of random sampling (efficiency ratio 7.98, 95% CI 6.23–9.74, P < 0.001; Fig. 2d–f) despite a more modest accuracy uplift (+3.1 percentage points, 95% CI 1.8–4.4, P = 0.003). S_3_ contributed a significant but narrower margin (+1.7 percentage points; Fig. 2g–i).

Boolean combinations of the three strategies traced a tunable trade-off between workload and safety (Table 1). The union of all strategies (S_1_ ∪S_2_ ∪S_3_) maximized error coverage at 81.6% while requiring a 40.7% review rate, whereas intersection rules (for example, S_1_ ∩ S_2_) pushed efficiency ratios above 9.0 but captured fewer than a third of errors. These results confirmed that combining orthogonal uncertainty signals provides a principled, adjustable mechanism for selective deferral in generative clinical AI.

**Table 1.**
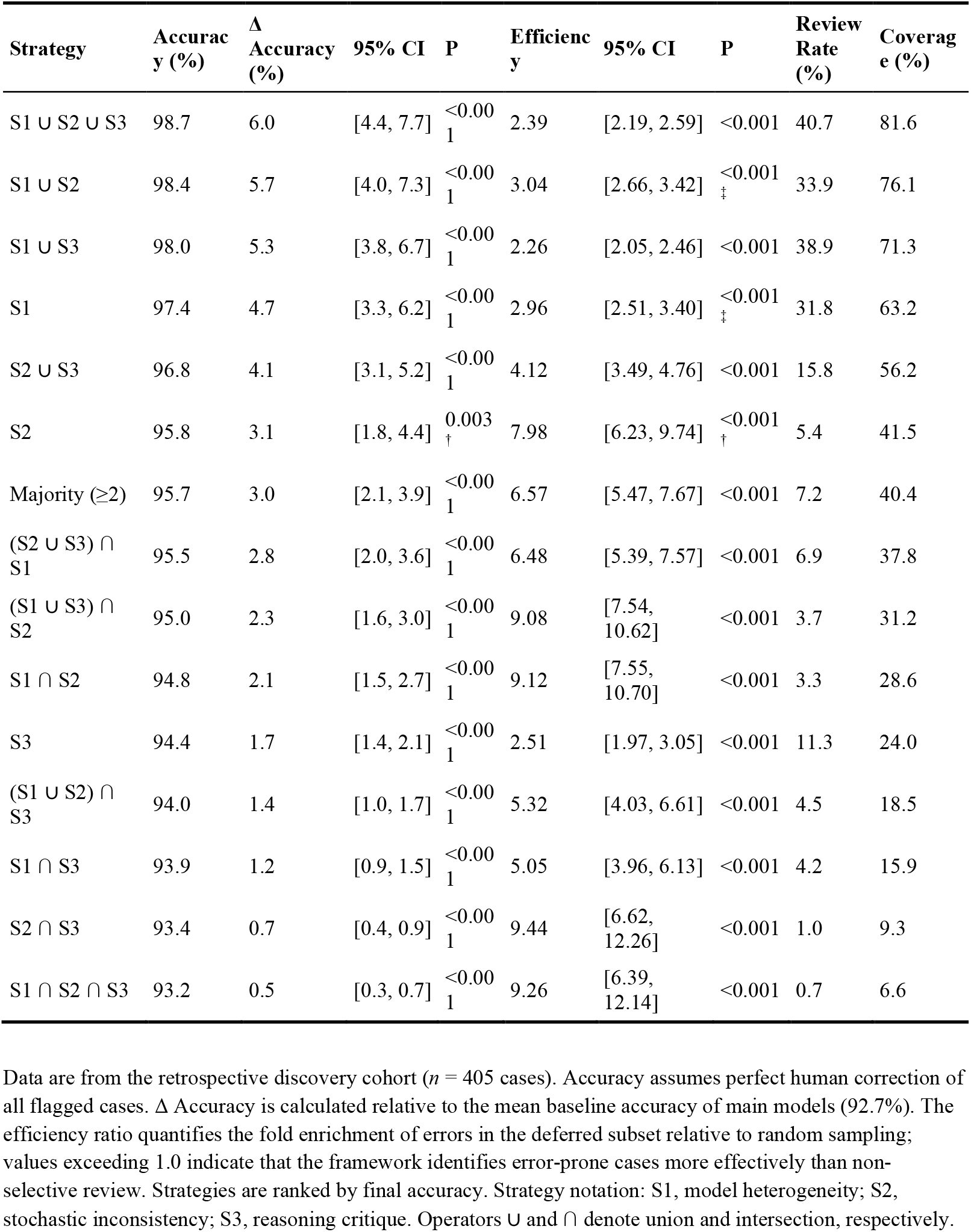
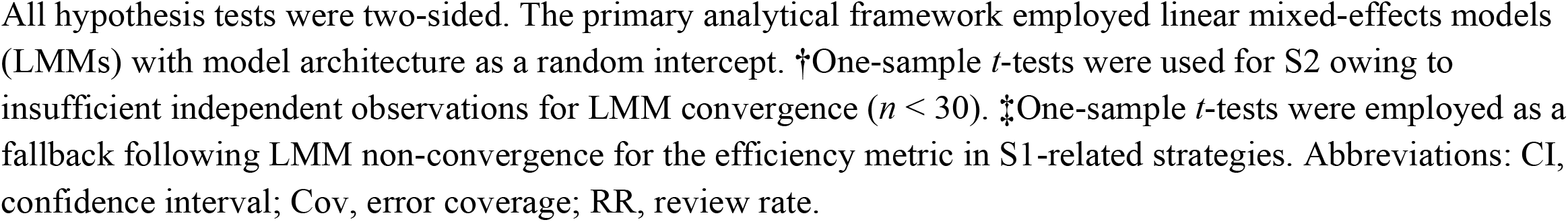
Comparative performance of uncertainty triangulation strategies and their Boolean combinations. Data are from the retrospective discovery cohort (*n* = 405 cases). Accuracy assumes perfect human correction of all flagged cases. Δ Accuracy is calculated relative to the mean baseline accuracy of main models (92.7%). The efficiency ratio quantifies the fold enrichment of errors in the deferred subset relative to random sampling; values exceeding 1.0 indicate that the framework identifies error-prone cases more effectively than non-selective review. Strategies are ranked by final accuracy. Strategy notation: S1, model heterogeneity; S2, stochastic inconsistency; S3, reasoning critique. Operators ∪and ∩ denote union and intersection, respectively. All hypothesis tests were two-sided. The primary analytical framework employed linear mixed-effects models (LMMs) with model architecture as a random intercept. †One-sample *t*-tests were used for S2 owing to insufficient independent observations for LMM convergence (*n* < 30). ‡One-sample *t*-tests were employed as a fallback following LMM non-convergence for the efficiency metric in S1-related strategies. Abbreviations: CI, confidence interval; Cov, error coverage; RR, review rate.

| Strategy | Accuracy (%) | $\Delta$ Accuracy (%) | 95% CI | P | Efficiency | 95% CI | P | Review Rate (%) | Coverage (%) |
| --- | --- | --- | --- | --- | --- | --- | --- | --- | --- |
| S1 ∪ S2 ∪ S3 | 98.7 | 6.0 | [4.4, 7.7] | $<0.001$ | 2.39 | [2.19, 2.59] | $<0.001$ | 40.7 | 81.6 |
| S1 ∪ S2 | 98.4 | 5.7 | [4.0, 7.3] | $<0.001$ | 3.04 | [2.66, 3.42] | $<0.001$ | 33.9 | 76.1 |
| S1 ∪ S3 | 98.0 | 5.3 | [3.8, 6.7] | $<0.001$ | 2.26 | [2.05, 2.46] | $<0.001$ | 38.9 | 71.3 |
| S1 | 97.4 | 4.7 | [3.3, 6.2] | $<0.001$ | 2.96 | [2.51, 3.40] | $<0.001$ | 31.8 | 63.2 |
| S2 ∪ S3 | 96.8 | 4.1 | [3.1, 5.2] | $<0.001$ | 4.12 | [3.49, 4.76] | $<0.001$ | 15.8 | 56.2 |
| S2 | 95.8 | 3.1 | [1.8, 4.4] | 0.003 | 7.98 | [6.23, 9.74] | $<0.001$ | 5.4 | 41.5 |
| Majority ($\geq 2$) | 95.7 | 3.0 | [2.1, 3.9] | $<0.001$ | 6.57 | [5.47, 7.67] | $<0.001$ | 7.2 | 40.4 |
| (S2 ∪ S3) ∩ S1 | 95.5 | 2.8 | [2.0, 3.6] | $<0.001$ | 6.48 | [5.39, 7.57] | $<0.001$ | 6.9 | 37.8 |
| (S1 ∪ S3) ∩ S2 | 95.0 | 2.3 | [1.6, 3.0] | $<0.001$ | 9.08 | [7.54, 10.62] | $<0.001$ | 3.7 | 31.2 |
| S1 ∩ S2 | 94.8 | 2.1 | [1.5, 2.7] | $<0.001$ | 9.12 | [7.55, 10.70] | $<0.001$ | 3.3 | 28.6 |
| S3 | 94.4 | 1.7 | [1.4, 2.1] | $<0.001$ | 2.51 | [1.97, 3.05] | $<0.001$ | 11.3 | 24.0 |
| (S1 ∪ S2) ∩ S3 | 94.0 | 1.4 | [1.0, 1.7] | $<0.001$ | 5.32 | [4.03, 6.61] | $<0.001$ | 4.5 | 18.5 |
| S1 ∩ S3 | 93.9 | 1.2 | [0.9, 1.5] | $<0.001$ | 5.05 | [3.96, 6.13] | $<0.001$ | 4.2 | 15.9 |
| S2 ∩ S3 | 93.4 | 0.7 | [0.4, 0.9] | $<0.001$ | 9.44 | [6.62, 12.26] | $<0.001$ | 1.0 | 9.3 |
| S1 ∩ S2 ∩ S3 | 93.2 | 0.5 | [0.3, 0.7] | $<0.001$ | 9.26 | [6.39, 12.14] | $<0.001$ | 0.7 | 6.6 |

### Pareto optimization and configuration selection

We next sought to identify a single operating point suitable for prospective deployment. Selecting DeepSeek-V3.1 with chain-of-thought (CoT) prompting as the main model (M_main_; standalone accuracy 92.4%), we performed a multi-objective Pareto optimization on the Discovery Cohort (n = 405), simultaneously minimizing physician review rate and maximizing final accuracy across all Boolean strategy combinations and model pairings (Fig. 3a). The all-union configuration (S_1_ ∪S_2_ ∪S_3_), using DeepSeek-V3.1 without optimized prompting as the auxiliary model and Qwen-32B as the checker, occupied the configuration that achieved the best balance of accuracy and workload reduction on the Pareto frontier. This configuration raised the model-specific accuracy from 92.4% to 100.0%, with 54.1% of cases routed to human review and an efficiency ratio of 1.9 (Fig. 3b,c). This configuration was locked for all subsequent validations, with no further tuning.

**Fig. 3.**
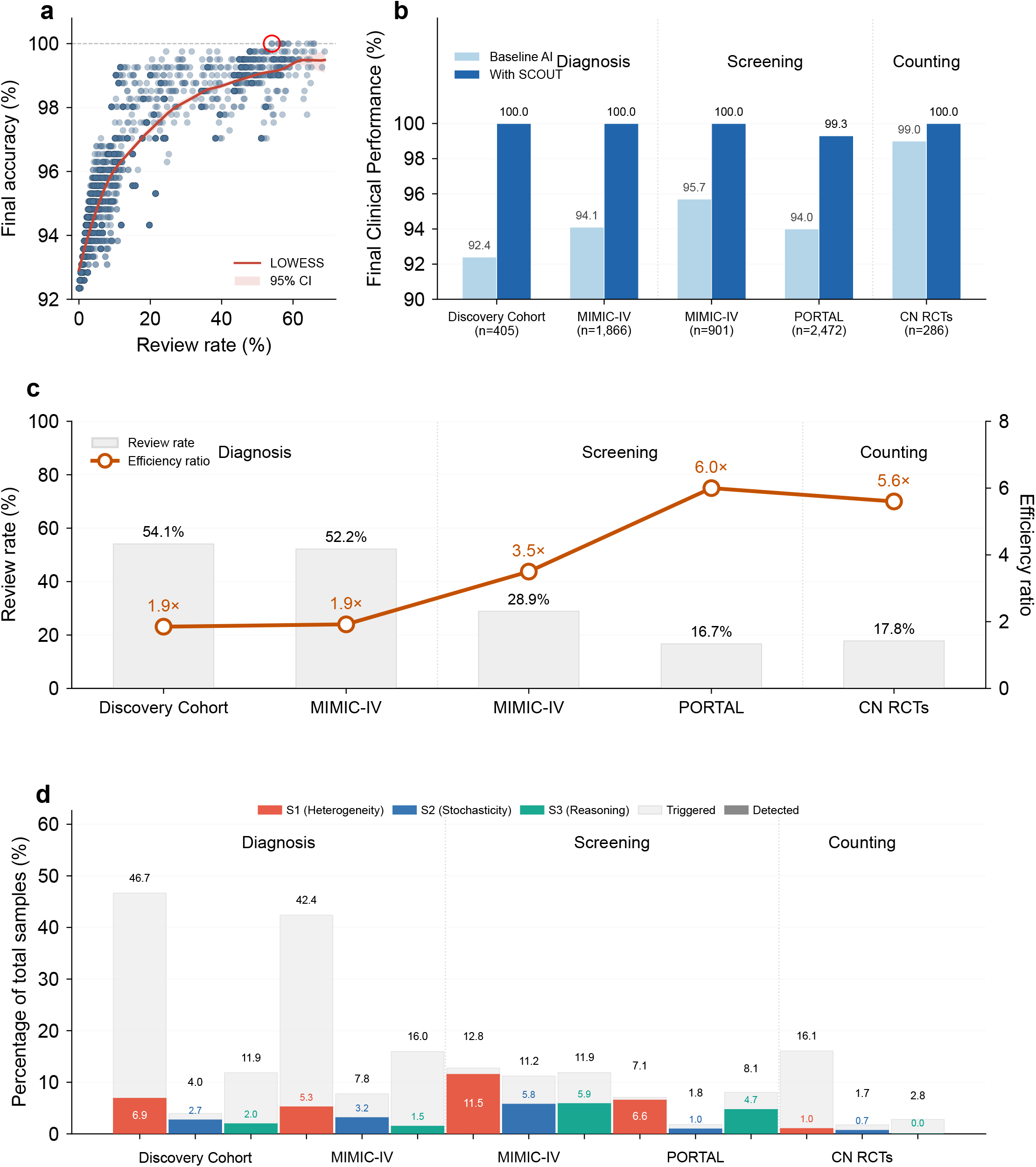
Generalizability of the SCOUT framework across clinical tasks and validation cohorts. **a**, Pareto efficiency frontier for the DeepSeek-V3.1 chain-of-thought model. Each data point represents a distinct operating point derived from varying Boolean combinations of the three verification strategies and auxiliary model pairings. The red curve indicates the LOWESS-fitted frontier (shaded area: 95% CI). The circled point marks the selected configuration (S1 ∪S2 ∪S3), which was locked for all subsequent validations. Data are from the discovery cohort (*n* = 405). **b**, Final clinical performance (accuracy for diagnostic and counting tasks; sensitivity for screening) comparing baseline AI (light blue) with the SCOUT-assisted workflow (dark blue) across six validation cohorts. Sample sizes are indicated in parentheses. **c**, Operational efficiency. Grey bars indicate the review rate (percentage of cases deferred to human verification); the orange line shows the efficiency ratio, quantifying the fold enrichment of errors in the deferred subset relative to random sampling. **d**, Contribution of individual verification strategies to case flagging and error detection. Light grey bars indicate the total proportion of cases flagged (trigger rate); coloured bars indicate the proportion of actual errors captured (detection yield). S1, model heterogeneity; S2, stochastic inconsistency; S3, reasoning critique. Abbreviations: CN, China; LOWESS, locally weighted scatterplot smoothing; MIMIC-IV, Medical Information Mart for Intensive Care version IV; RCTs, randomized clinical trials.

### Generalizability across clinical tasks and populations

We assessed the locked SCOUT configuration across three clinically distinct tasks and four independent cohorts (Fig. 3b–d).

For the diagnostic reasoning task (coronary heart disease subtyping), the framework was applied to the MIMIC-IV dataset, a cross-lingual challenge involving English-language US critical care records (Extended Data Table 2). SCOUT raised accuracy from 94.1% to 100% while restricting human review to 52.2% of cases (efficiency ratio 1.9).

For the cancer screening task (liver cancer risk detection from radiology reports), the clinical stakes demanded near-zero tolerance for missed malignancies. In the MIMIC-IV cohort (n = 901), SCOUT achieved both 100% sensitivity and 100% specificity (Table 2). In the PORTAL cohort (n = 2,472), comprising cirrhotic patients from two Chinese tertiary centres (Extended Data Table 3), the system attained 99.3% sensitivity and 100% specificity, reducing the review volume by more than 80% while maintaining a near-zero-miss safety profile.

**Table 2.** External validation of SCOUT for liver cancer screening. Data represent performance across two independent multicentre imaging screening cohorts. Abbreviations: Sens., sensitivity; Spec., specificity. All values are percentages.

| Dataset | Standalone AI |  | SCOUT |  |
| --- | --- | --- | --- | --- |
|  | Sens. | Spec. | Sens. | Spec. |
| PORTAL Cohort | 94.0 | 90.9 | 99.3 | 100.0 |
| MIMIC-IV Cohort | 95.7 | 84.9 | 100.0 | 100.0 |

For the information extraction task (counting diseased coronary vessels from angiographic reports), a task requiring precise numerical reasoning across heterogeneous report formats (Extended Data Table 4), SCOUT achieved 100% accuracy with a 5.6-fold efficiency ratio. Across all cohorts, the framework enriched errors in the deferred subset at 1.9 to 6.0 times the random-sampling rate (Fig. 3c), indicating that the meta-verification logic transfers across tasks without retraining.

One silent failure case was identified in which all three verification strategies failed to flag an incorrect prediction (Extended Data Table 5). This case originated from intrinsic ambiguity in the source text rather than from a model reasoning error. The CT report described only diffuse low-density foci without morphological detail sufficient to distinguish regenerative nodules from malignancy, a distinction that requires original imaging review. This case delineates a boundary condition in which the clinical text itself cannot support a definitive classification; model-level triangulation shares the same information ceiling. Complementary safeguards for such scenarios are discussed below.

## Discussion

We report the retrospective development and external validation of SCOUT, to our knowledge the first model-agnostic meta-verification framework designed specifically for generative AI oversight in clinical medicine. Extensive external validation across three distinct clinical tasks, spanning diagnostic reasoning, imaging screening and logical information extraction, demonstrated that the framework generalizes without task-specific retraining. These findings establish SCOUT as a practical audit infrastructure for scaling clinical AI deployment.

SCOUT addresses fundamental limitations of two established oversight paradigms^23,24^. Universal clinician-in-the-loop review, while proven in mammography screening^25^ and sepsis management^8^, becomes unscalable when applied to the high-throughput outputs of large language models. Learning-to-defer approaches offer a selective alternative: CoDoC^15^ achieved a landmark reduction in reader workload by up to 66% in mammography while surpassing the double-reading standard. However, that paradigm requires access to scalar confidence scores and task-specific calibration data, posing two barriers for generative AI. First, large language models produce free-text sequences rather than calibrated probabilities^9,17,26^. Second, the need for labeled tuning data limits deployability in resource-constrained settings^14,22,27,28^. These architectural constraints are compounded by evidence that threshold-based abstention can paradoxically alter clinician error patterns, promoting under-diagnosis^21^. By triangulating external, model-agnostic uncertainty signals, SCOUT eliminates the dependency on internal confidence or task-specific training, aligning with the high-sensitivity demands of clinical decision support.

The framework derives its robustness from three complementary verification strategies, each targeting a distinct failure mode. Model heterogeneity (Strategy 1) introduces architectural diversity to break the systematic reasoning errors to which any single model is prone^6,29^, functioning as an algorithmic analogue of independent double reading in radiology. Stochastic Inconsistency (Strategy 2) leverages generative non-determinism as a proxy for decision-boundary uncertainty^30^. When repeated sampling produces divergent outputs, the prediction is likely brittle. This proved to be the most cost-effective signal, achieving an efficiency ratio of 7.98. Reasoning Critique (Strategy 3) deploys an external checker model to audit the chain-of-thought logic of the primary model. Although this strategy contributed a statistically significant supplementary safety layer, it was the least effective individually, highlighting the well-documented risk that well-aligned language models can generate fluent yet incorrect rationales^6,20^. Collectively, these results indicate that outcome-oriented triangulation (Strategies 1 and 2) provides a stronger safety guarantee than process-oriented verification alone.

This study has several limitations. Validation was confined to text-based clinical records, and applicability to multimodal data remains to be established. The efficiency of the framework is contingent upon a sufficiently accurate base model; poor baseline performance would trigger frequent deferrals and erode operational gains. We observed one silent failure case in which all verification layers failed to trigger, marked by an ambiguous clinical description lacking key discriminative details. This represents a fundamental boundary condition: when multiple models share systematic misconceptions about inherently ambiguous inputs, triangulation alone cannot guarantee detection. For scenarios prone to this failure mode, the framework should be complemented by rule-based filtering or targeted review of predefined high-risk categories. As multimodal AI systems increasingly combine imaging with generative reasoning, future hybrid architectures could integrate confidence-based deferral for image interpretation^15^ with model-agnostic triangulation for text generation, achieving complementary safety coverage across modalities. Importantly, the present study is limited to retrospective data, and all performance projections assume perfect human correction of flagged cases— an assumption that must be tested in real-world workflows. A prospective, randomized controlled trial (registration in progress) is planned to evaluate SCOUT’s impact on diagnostic accuracy, physician workload, and clinical decision-making under routine conditions.

In summary, this retrospective study establishes SCOUT as a promising, scalable blueprint for responsible AI deployment in medicine. By decoupling automation speed from safety requirements through an external meta-verification layer, the framework offers a pragmatic path to democratize expert-level diagnostics while keeping the human meaningfully in the loop. Prospective validation is the necessary next step to translate these findings into clinical practice.

## Methods

### The SCOUT Framework

We designed SCOUT as a model-agnostic audit layer that routes only high-risk AI outputs to clinicians while allowing low-risk predictions to be accepted automatically. Let *x* denote a clinical case and *y* the ground-truth label. The main model *M*_main_ generates an initial prediction *ŷ* = *M*_main_(*x*). SCOUT introduces a binary deferral function *D*(*x*) that partitions all cases into two streams:

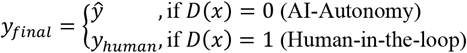

The deferral function is constructed from the Boolean combination of three orthogonal verification strategies (*S*_1_, *S*_2_, *S*_3_), each targeting a distinct failure mode of large language models. The specific Boolean operator was optimized via Pareto search on the Discovery Cohort (see below).

### Uncertainty triangulation strategies

#### Strategy 1 model heterogeneity

A single model can harbour systematic reasoning blind spots, analogous to the echo-chamber effect in clinical second opinions^6,29^. To break such correlated errors, we introduced an auxiliary model *M*_aux_ whose architecture, parameter scale, inference mode, or prompt design differed substantially from *M*_main_. The flag *S*_1_(*x*) = 1 is triggered whenever the two models disagree: *M*_main_(*x*) ≠ *M*_aux_(*x*). Prompt heterogeneity was achieved by contrasting a baseline prompt (task definitions and format requirements only) with an optimized prompt augmented by guideline-derived clinical knowledge, few-shot exemplars, explicit reasoning directives, and negative constraints designed to pre-empt common error patterns.

#### Strategy 2 stochastic inconsistency

When a model sits near a decision boundary, repeated sampling under identical conditions can yield divergent outputs, much as a hesitating clinician might waver between two diagnoses^30^. We exploited this property by executing one additional independent inference run of *M*_main_ at the same temperature setting. The flag *S*_2_(*x*) = 1 is activated if the re-sampled output contradicts the initial prediction, thereby exposing instability in the model’s decision confidence without requiring access to internal logits^17^.

#### Strategy 3 reasoning critique

A correct final answer can arise from flawed reasoning, a lucky guess that may not generalize^6^. To detect such cases, we deployed a checker model *M*_check_ to audit the chain-of-thought (CoT) trace produced by *M*_main_. The checker performs a binary pass/fail judgment targeting logical fallacies within the reasoning trace. The flag *S*_3_(*x*) = 1 is triggered if the verdict is fail.

### Configuration selection and model implementation

The optimal SCOUT configuration was identified through multi-objective Pareto optimization on the retrospective Discovery Cohort. The search space encompassed all boolean combinations of the three strategies together with the candidate model architectures for each verification role. The two competing objectives were to minimize the review rate (physician workload) and maximize the final accuracy (patient safety).

We systematically evaluated 22 configurations across a suite of foundational models, including GPT-5.1, DeepSeek-V3.1/V3.2, and Qwen3 (235B/32B/8B). To mirror the performance level of models already deployed in clinical practice, only those achieving a baseline accuracy of 90% or above were considered as candidates for *M*_main_. Open-source models were accessed through the Alibaba Cloud API (https://bailian.console.aliyun.com), and GPT-5.1 was accessed through the OpenAI API (https://platform.openai.com). To balance reproducibility with the stochastic diversity needed for uncertainty estimation, hyperparameters were controlled as follows: top_p was fixed at 0.95 for all runs; for standard (non-reasoning) inference, temperature was set to 0.1; for chain-of-thought reasoning, temperature followed each provider’s official recommendation (0.6 for DeepSeek-V3.1 and Qwen3; 1.0 for DeepSeek-V3.2). GPT-5.1 parameters were managed through API defaults.

### Data sources and study population

#### Discovery Cohort (model derivation)

To derive the SCOUT parameters, we assembled 405 high-quality admission records from two randomized clinical trials in China (Extended Data Fig.1a): AIM-CHD^32^ (NCT06686056) and SMART-CHD (NCT07031531). Eligible patients were adults (≥18 years) with suspected coronary heart disease and available unstructured admission notes. Cases with insufficient documentation (fewer than 50 words) were excluded.

#### External Validation Cohorts (Multi-Task Evaluation)

Generalizability was assessed across three clinical tasks representing distinct cognitive modalities.

#### Task 1: complex diagnostic reasoning (CHD subtyping)

Cross-lingual validation was performed on the US-based MIMIC-IV database (Extended Data Fig.1a). From 107,553 admissions, we identified cardiothoracic cases and drew a random sample of 2,000 instances. After excluding non-primary CHD admissions, 1,866 cases were retained. The task required classifying each patient as ST-elevation myocardial infarction (STEMI), non-ST-elevation myocardial infarction (NSTEMI), unstable angina or chronic coronary syndromes on the basis of discharge summaries.

#### Task 2: large-scale CT screening (liver cancer risk detection)

Cross-task validation was conducted for liver cancer risk detection from radiology reports (Extended Data Fig.1b). A cohort of 3,373 cirrhotic patients was aggregated from the PORTAL cohort (China, cases from 2 hospitals) and MIMIC-IV-Radiology (US). After excluding duplicates, patients with a prior history of malignancy and non-diagnostic scans, 2,472 Chinese and 901 US cases were retained for analysis.

#### Task 3: logical information extraction (vessel counting)

This task evaluated high-precision reasoning by requiring quantification of coronary stenosis severity (categorized as 0-, 1-, 2- or 3-vessel disease) from angiographic reports. A held-out set of 286 cases was sampled from the AIM-CHD and SMART-CHD trials (Extended Data Fig.1c). Severe stenosis was defined as ≥50% diameter reduction for the left main artery and ≥70% for non-left-main arteries.

### Gold-standard adjudication

#### Cardiovascular tasks (Tasks 1 and 3)

Ground-truth labels were established in accordance with the 2024 ESC Guidelines for the management of chronic coronary syndromes and the 2025 AHA Guidelines for acute coronary syndromes. Adjudication followed a double-blinded consensus workflow: two board-certified cardiologists from Fuwai Hospital independently reviewed the raw medical records, and discrepancies were resolved by a senior consultant with over 25 years of experience. All adjudicators were blinded to model predictions and received dedicated training on CHD subtyping before labelling.

#### Hepatology task (Task 2)

Standard radiology reports comprise two sections: *findings* (descriptive observations) and *impression* (the radiologist’s diagnostic conclusion). We derived the primary ground truth from the impression section of the original clinical reports, each of which had undergone standard dual-signature verification. To harmonize labels across institutions, we applied a semi-automated pipeline: an ancillary LLM first structured the impressions into a four-tier risk scale (confirmed liver cancer or high risk versus benign or no lesion), which was then binarized into high-risk and low-risk categories. An attending hepatologist manually adjudicated all instances of discordance between model predictions and pipeline-derived labels, thereby ruling out information asymmetry or original reporting errors as confounders.

### Statistical analysis

#### Evaluation metrics

Four metrics were defined to quantify the clinical utility and cost-effectiveness of the framework. The *review rate* (RR) is the proportion of the total cohort deferred to human verification: for diagnostic tasks (Tasks 1 and 3),

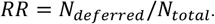

For the screening task (Task 2), the review scope was expanded to include both SCOUT-flagged cases and all model-predicted positives (*D*(*x*) = 1 ∪*ŷ* = 1) to ensure verification of all potential pathologies regardless of algorithmic uncertainty.

The *error coverage* (EC) is the proportion of initial model errors intercepted by the review mechanism:

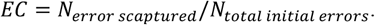

The *efficiency ratio* (ER) quantifies the density of errors within the deferred subset relative to random sampling:

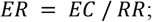

a value exceeding 1.0 indicates that the framework identifies error-prone cases more effectively than non-selective full review.

*Final clinical performance* was projected assuming expert reviewers correctly resolve all flagged cases. For classification tasks:

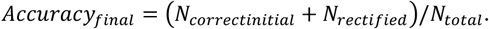

For the screening task, owing to class imbalance and the greater clinical value of avoiding missed diagnoses, we reported post-review sensitivity.

#### Algorithmic meta-verification analysis

For retrospective evaluations, we used linear mixed-effects models to account for the hierarchical data structure in which observations were nested within model architectures. Verification strategies were treated as fixed effects and model architectures as random intercepts, yielding adjusted estimates of the efficiency ratio and accuracy improvement. For Strategy 2, where variability arose solely from stochastic sampling within a single architecture rather than from nested auxiliary components, significance was assessed with a one-sample *t*-test.

### Ethics and Regulatory Compliance

The study protocol complied with the Declaration of Helsinki and was approved by the Institutional Review Boards of Fuwai Hospital, and Peking University First Hospital. Access to the MIMIC-IV database^33^ was granted under the PhysioNet Data Use Agreement following completion of CITI Program ethical training. Given the retrospective nature of the model development and external validation phases, individual informed consent was waived by the respective IRBs owing to minimal risk and the exclusive use of de-identified data.

## Data Availability

All data produced in the present study are available upon reasonable request to the authors

## Data availability

The MIMIC-IV database (https://physionet.org/content/mimiciv/3.1/) is freely available to investigators who establish a PhysioNet account and satisfy the required data access protocols. The AIM-CHD, SMART-CHD, and PORTAL datasets are available from the corresponding author upon reasonable request.

## Acknowledgements

This work was supported by CAMS Innovation Fund for Medical Sciences (CIFMS) (2024-I2M-C&T-B-040), Noncommunicable Chronic Diseases-National Science and Technology Major Project (2023ZD0504005), CAMS Innovation Fund for Medical Sciences (CIFMS) (2024-I2M-ZH-004), and the National High Level Hospital Clinical Research Funding (2025-GSP-GG-18-3, 2025-GSP-GG-14).

## Author contributions

Y.W., X.G. and G.W. contributed to the conception and design of the study; Z.B. and M.H. contributed to the data analysis and writing of the manuscript; H.H. and X.D. provided technical assistance; Q.F., J.L.(Jingrong Lai), R.Z., M.L.(Mengyuan Liu), Z.W., X.W.(Ximei Wang), S.Z., Y.Z., H.C., Y.Q., Q.S., J.X., F.H., X.L., H.C.(Hongwu Chen), M.Z., B.X., J.L.(Jiabao Liu), and N.G. contributed to the data acquisition, curation and analysis. All authors contributed to the drafting and revising of the paper.

## EXTENDED DATA FIGURES

**Extended Data Figure 1.**
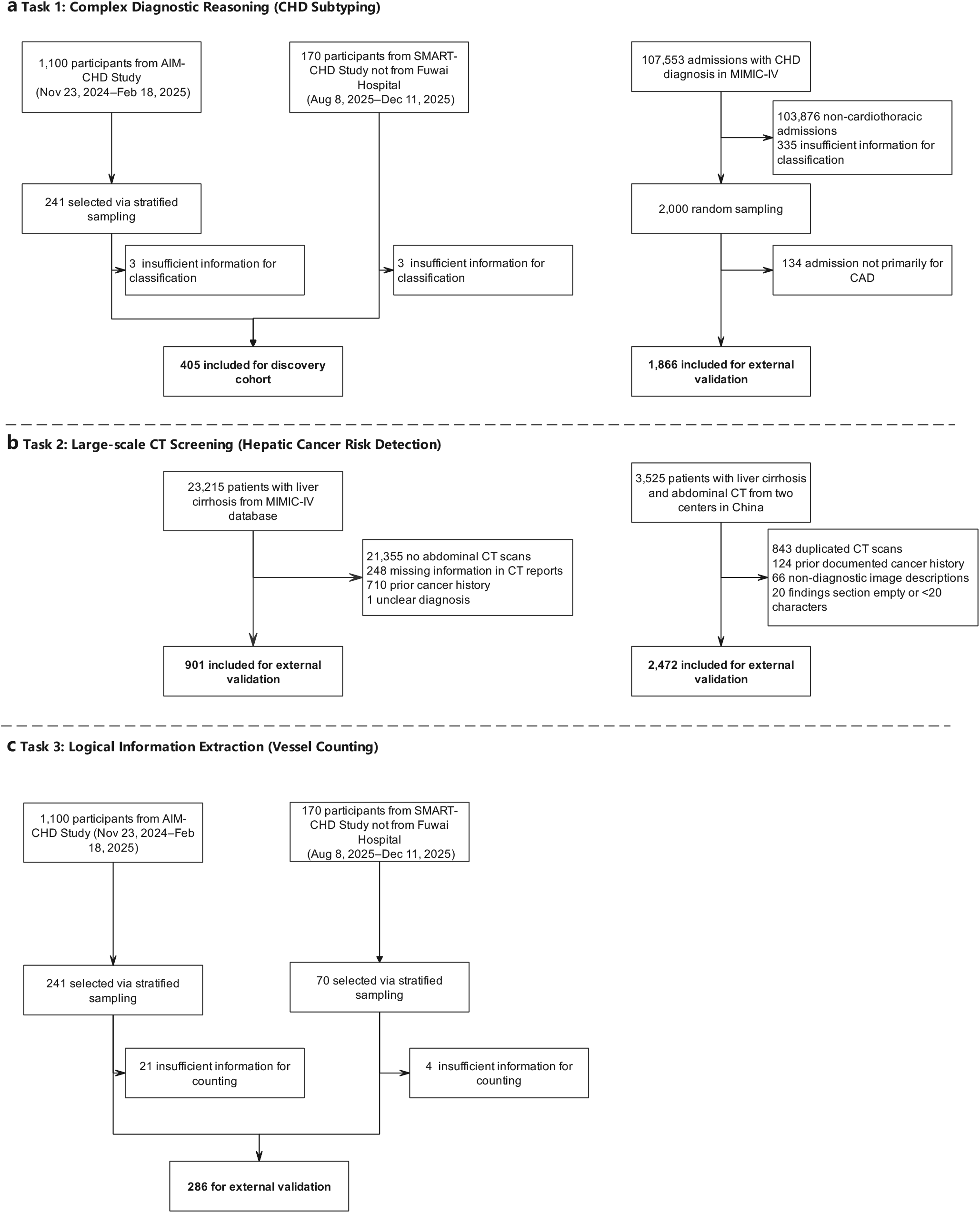
Study flowcharts for dataset construction and participant enrolment. **a**, Task 1: complex diagnostic reasoning (coronary heart disease subtyping). Cases were derived from the AIM-CHD and SMART-CHD trials (discovery cohort, *n* = 405) and from the MIMIC-IV database (external validation, *n* = 1,866). **b**, Task 2: large-scale CT screening (liver cancer risk detection). Cirrhotic patients were assembled from two Chinese centres (*n* = 2,472) and from MIMIC-IV (*n* = 901). **c**, Task 3: logical information extraction (vessel counting). A held-out set of 286 cases was sampled from the AIM-CHD and SMART-CHD trials.

## EXTENDED DATA TABLES

**Extended Data Table 1.**
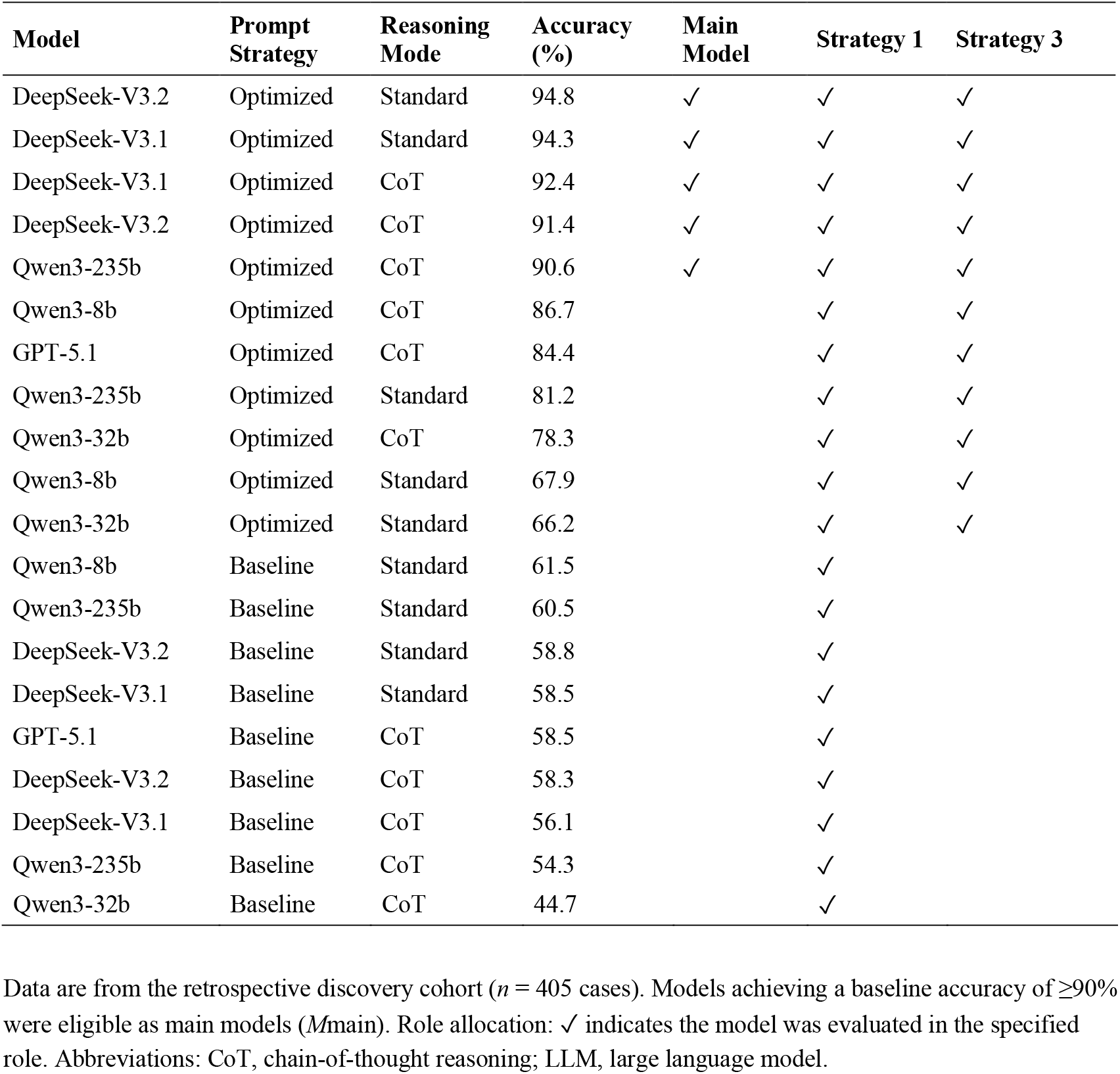
Performance of candidate large language models and their role allocation in the SCOUT framework. Data are from the retrospective discovery cohort (*n* = 405 cases). Models achieving a baseline accuracy of ≥90% were eligible as main models (*M*main). Role allocation: ✓ indicates the model was evaluated in the specified role. Abbreviations: CoT, chain-of-thought reasoning; LLM, large language model.

**Extended Data Table 2.**
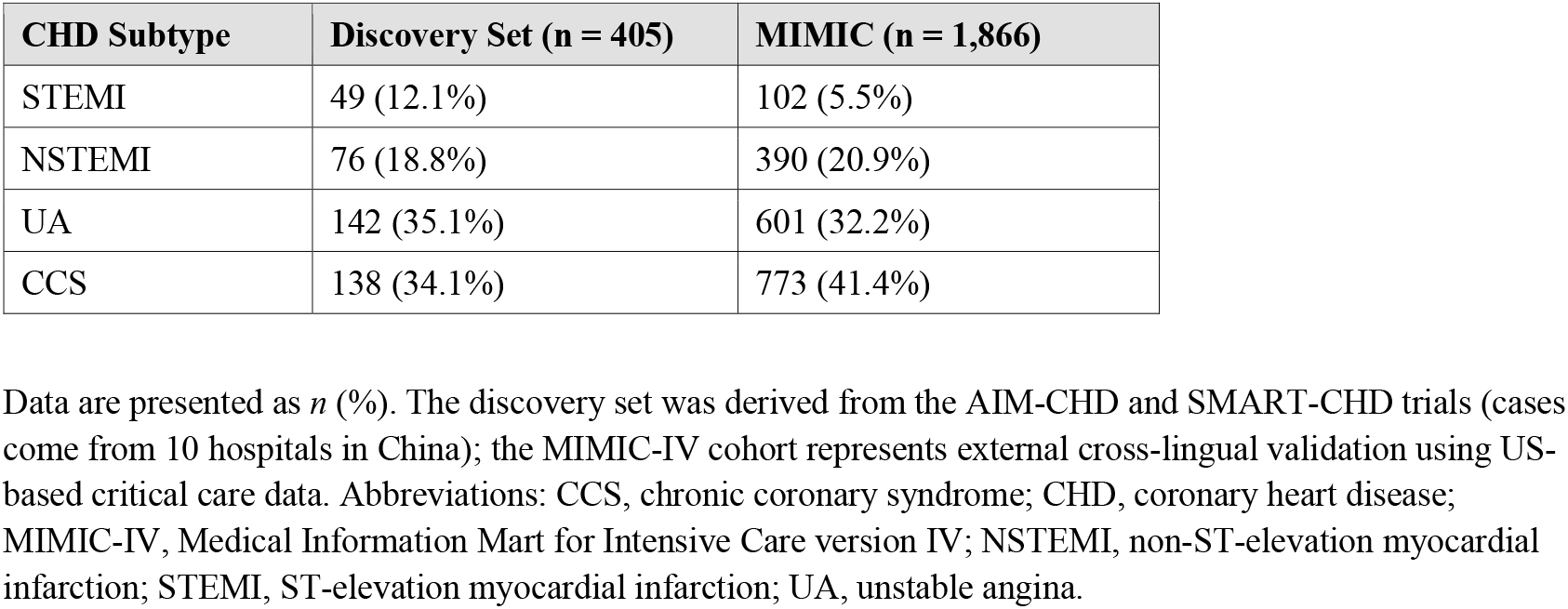
Distribution of coronary heart disease subtypes across study cohorts. Data are presented as *n* (%). The discovery set was derived from the AIM-CHD and SMART-CHD trials (cases come from 10 hospitals in China); the MIMIC-IV cohort represents external cross-lingual validation using US-based critical care data. Abbreviations: CCS, chronic coronary syndrome; CHD, coronary heart disease; MIMIC-IV, Medical Information Mart for Intensive Care version IV; NSTEMI, non-ST-elevation myocardial infarction; STEMI, ST-elevation myocardial infarction; UA, unstable angina.

**Extended Data Table 3.** Distribution of liver lesion risk categories in the liver cancer screening cohort. Data are presented as *n* (%). The Chinese cohort included patients from Peking University First Hospital and the Fifth People’s Hospital of Wuxi. For binary classification, low-risk includes ‘no focal lesion’ and ‘benign lesion’; high-risk includes ‘high-risk lesion’ and ‘confirmed liver cancer’. Abbreviations: MIMIC-IV, Medical Information Mart for Intensive Care version IV.

| Lesion Category | MIMIC-IV Cohort | PORTAL Cohort |
| --- | --- | --- |
| | $n$ (%) | $n$ (%) |
| No focal lesion | 846 (93.9) | 2,048 (82.8) |
| Benign lesion | 9 (1.0) | 290 (11.7) |
| High-risk lesion | 44 (4.9) | 118 (4.8) |
| Confirmed liver cancer | 2 (0.2) | 16 (0.6) |
| **Total** | **901 (100.0)** | **2,472 (100.0)** |

**Extended Data Table 4.** Distribution of significant coronary artery disease by number of vessels involved. Data (*n* = 286) represent a subset from the AIM-CHD and SMART-CHD trials. Significant stenosis was defined as ≥50% luminal diameter reduction for the left main artery and ≥70% for other epicardial arteries.

| Number of Vessels with Significant Lesions | n | % |
| --- | --- | --- |
| 0 | 16 | 5.6 |
| 1 | 100 | 35.0 |
| 2 | 93 | 32.5 |
| 3 | 77 | 26.9 |
| Total | 286 | 100.0 |

Data ( $n = 286$ ) represent a subset from the AIM-CHD and SMART-CHD trials. Significant stenosis was defined as $\geq 50\%$ luminal diameter reduction for the left main artery and $\geq 70\%$ for other epicardial arteries.

**Extended Data Table 5.**
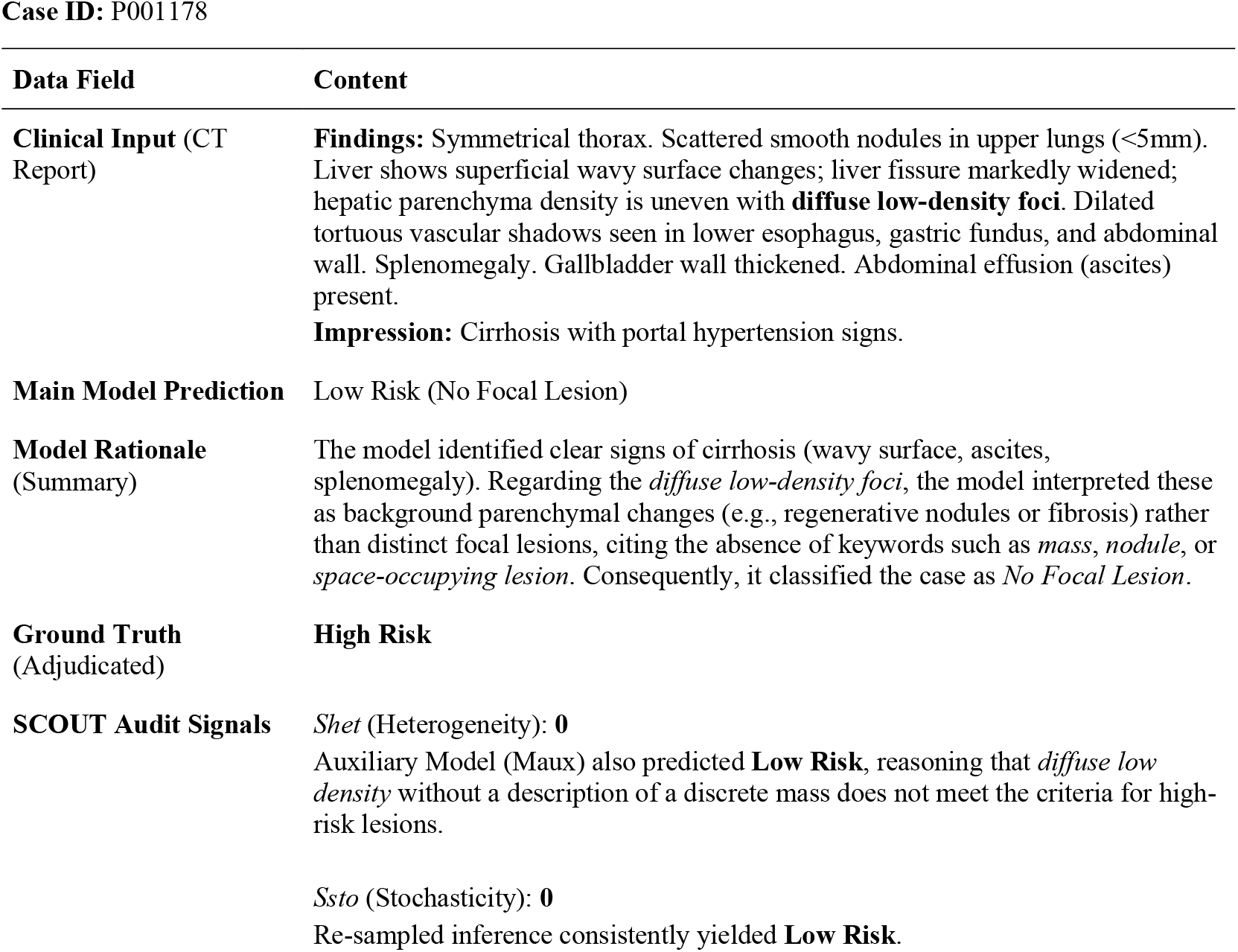
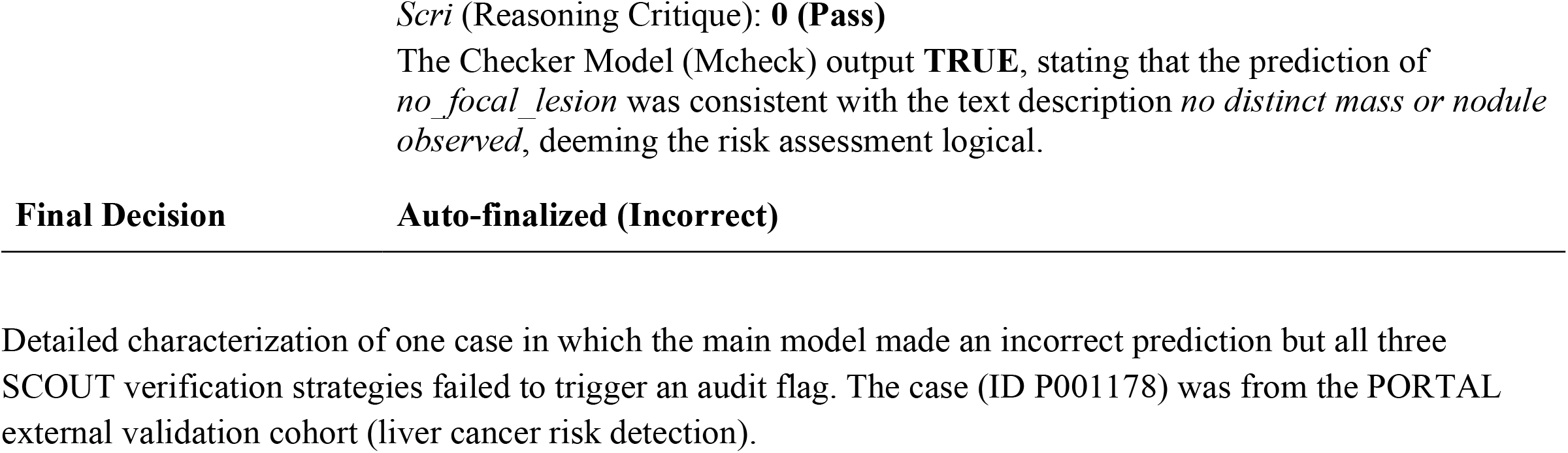
Characterization of representative silent failure case. **Case ID:** P001178 Detailed characterization of one case in which the main model made an incorrect prediction but all three SCOUT verification strategies failed to trigger an audit flag. The case (ID P001178) was from the PORTAL external validation cohort (liver cancer risk detection).

